# Heart Rate, Electrocardiographic Subclinical Myocardial Injury, and Long-Term Mortality

**DOI:** 10.64898/2026.02.27.26347281

**Authors:** Patrick Cheon, Mohamed A. Mostafa, Mai Z Soliman, Richard Kazibwe, Elsayed Z Soliman

## Abstract

**Background:** Elevated resting heart rate is associated with increased mortality, but the underlying mechanisms remain incompletely understood. Subclinical myocardial injury (SCMI), defined by a Cardiac Infarction/Injury Score (CIIS) ≥10, represents silent cardiac damage that predicts poor cardiovascular (CV) outcomes and may partially explain this association.

**Methods:** We analyzed 7,152 participants from NHANES III who underwent ECG recording and were free of cardiovascular disease. Heart rate was categorized as bradycardia (≤50 bpm), normal (>50–<100 bpm), or tachycardia (≥100 bpm). Mortality was assessed through National Death Index linkage. Logistic and Cox regression models evaluated associations with SCMI and mortality, respectively, and attenuation was assessed by change in hazard ratios after adjusting for SCMI.

**Results:** SCMI was present in 1,744 (24.3%) participants. Tachycardia was associated with increased odds of SCMI (adjusted OR 2.34, 95% CI 1.42–3.88). Over 13.9 years median follow-up, 2,311 (32.3%) died from all causes and 933 (13.1%) from CV causes. Tachycardia was associated with increased all-cause mortality (HR 3.58, 95% CI 2.63–4.88) and CV mortality (HR 2.05, 95% CI 1.06–3.79). Adjustment for SCMI attenuated the tachycardia–CV mortality association by 8.6% and all-cause mortality by 5%. Bradycardia was not associated with SCMI or mortality.

**Conclusion:** These findings suggest that SCMI partially mediates the heart rate–mortality relationship and that ECG-based assessment of SCMI may enhance risk stratification in individuals with elevated resting heart rate.

## Introduction

Elevated resting heart rate is associated with increased risk of all-cause and cardiovascular (CV) mortality across diverse populations ^1–4^. This association persists after adjustment for traditional CV risk factors, suggesting that elevated heart rate may contribute to underlying pathophysiological processes beyond its role as a simple vital sign. Proposed mechanisms include increased myocardial oxygen demand, reduced diastolic perfusion time, and autonomic imbalance ^5,6^. However, these mechanisms do not fully account for the excess mortality risk associated with elevated heart rate, and the precise pathways linking elevated heart rate to adverse outcomes remain incompletely understood.

Subclinical myocardial injury (SCMI) represents silent cardiac damage occurring in the absence of overt clinical symptoms. SCMI can be detected using the Cardiac Infarction/Injury Score (CIIS), a validated electrocardiographic measure, and is strongly associated with future CV events and mortality ^7–9^. Given that elevated resting heart rate increases myocardial oxygen demand, SCMI may represent a plausible mechanistic pathway through which elevated heart rate contributes to mortality ^5,6^. However, the relationship between resting heart rate and SCMI has not been systematically evaluated, and whether SCMI partially explains the association between elevated heart rate and mortality remains unknown.

We hypothesized that higher resting heart rate is associated with SCMI and that SCMI partially accounts for the association between resting heart rate and long-term all-cause and cardiovascular mortality. Using data from the United States Third National Health and Nutrition Examination Survey (NHANES III), we evaluated the association between resting heart rate and SCMI as well as examined whether SCMI partially explains the relationship between elevated resting heart rate and long-term all-cause and CV mortality.

## Methods

### Study Population

NHANES III is a program designed to assess health and nutritional status among U.S. adults and children living in the community. It is conducted by the National Center for Health Statistics (NCHS), part of the Centers for Disease Control and Prevention. NHANES III was carried out between 1988 and 1994. All participants gave written informed consent, and the study protocol was approved by the NCHS institutional review board. Further information on the survey’s design, methodology, and data availability has been documented in prior publications ^10^.

For this analysis, we included NHANES III participants with available electrocardiogram (ECG) data at baseline. By design, only NHANES III participants aged 40 years or older underwent ECG recording. We excluded those with prior cardiovascular disease (CVD) (prior myocardial infarction, heart failure, or stroke), those not in sinus rhythm, those using antiarrhythmic medications, and those with missing key variables needed for analysis. After all exclusions, 7,152 participants were included in the analysis **(Figure 1)**.

**Figure 1.**
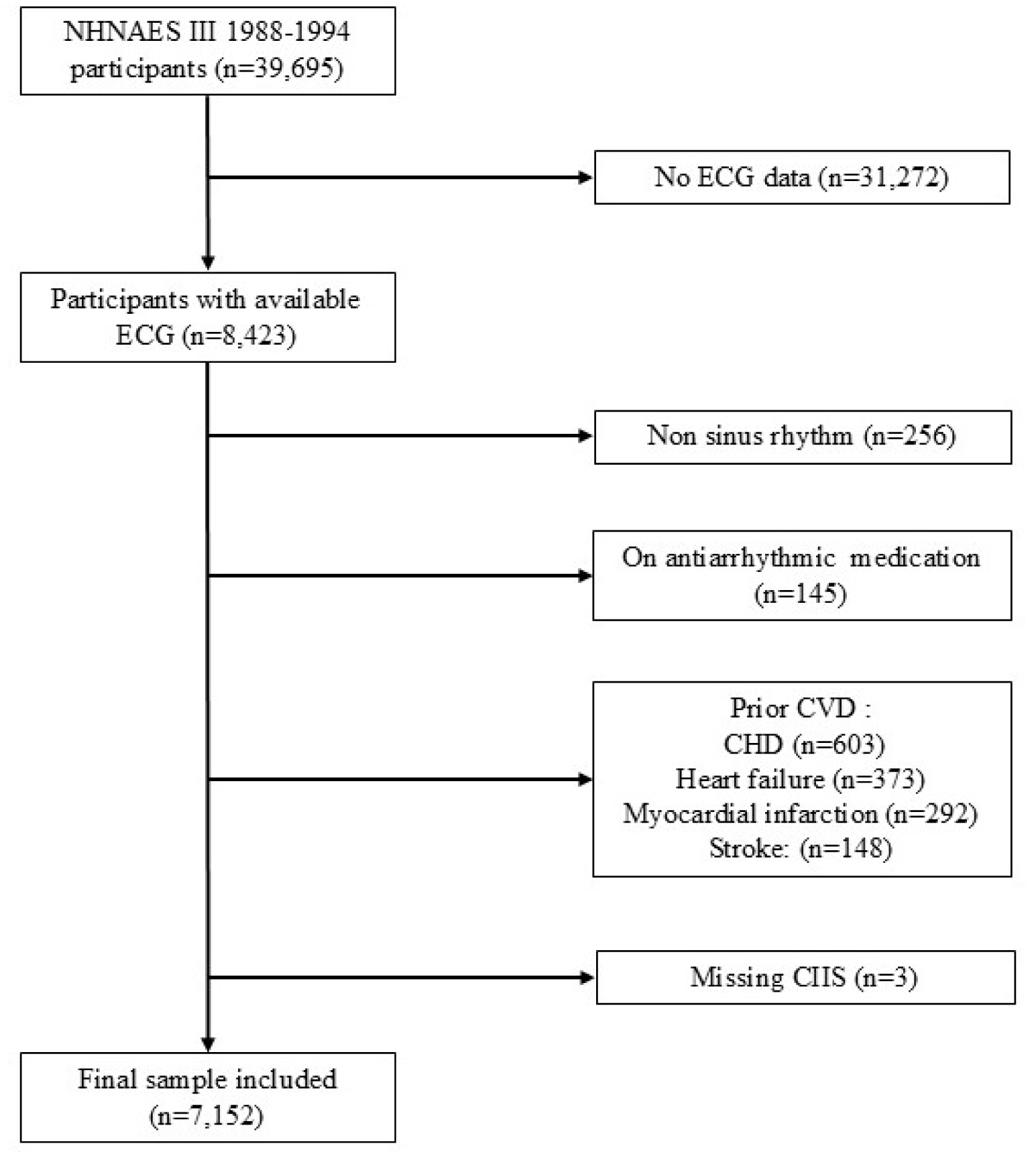
Study population flow diagram. Abbreviations: ECG, electrocardiogram; CVD, cardiovascular disease; CHD, coronary heart disease; CIIS, cardiac infarction/injury score.

### Ascertainment of Resting Heart Rate

In NHANES III, a resting 12-lead ECG was obtained using a Marquette MAC 12 electrocardiograph (Marquette Medical Systems, Milwaukee, WI, USA) during a physical examination conducted within a mobile examination center. Resting heart rate was derived from the standard 12-lead ECG recording. For the primary analysis, participants were categorized into three groups based on resting heart rate: bradycardia (≤50 beats per minute [bpm]), normal heart rate (>50 to <100 bpm), and tachycardia (≥100 bpm). In secondary analyses, heart rate was also modeled as a continuous variable per 10-bpm increase.

### Defining Subclinical Myocardial Injury (SCMI)

The electrocardiographic CIIS was used to define SCMI. The digital signals of the ECG tracings were sent to the Epidemiological Cardiology Research Center (EPICARE) of Wake Forest University School of Medicine (Winston-Salem, NC, USA) for central processing. ECGs underwent visual inspection by skilled technicians before being automatically processed using the GE 12-SL program (Marquette Medical Systems, Milwaukee, WI, USA).

The CIIS is a weighted scoring system designed to assess the likelihood of myocardial injury or ischemia based on quantitative ECG features. It incorporates a total of 15 components, including 11 discrete and 4 continuous ECG variables, derived from standard 12-lead ECGs.

These elements capture abnormalities in Q waves, R waves, T waves, and the ST segment, and together provide a risk-stratified assessment of cardiac injury. The CIIS is structured to allow for both manual interpretation and automated processing, making it suitable for use in large population-based studies ^7,8^.

In the NHANES III dataset, CIIS values were initially stored after being multiplied by 10 to avoid the use of decimal points in the database. For the purpose of this analysis, the values were converted back to their original scale by dividing by 10. In accordance with established definitions in the literature, a CIIS of 10 or greater was used to define the presence of SCMI ^11–15^.

### Ascertainment of Mortality

The primary outcomes were all-cause and CV mortality, determined through linkage to the National Death Index with follow-up through 31 December 2015. Cardiovascular deaths were identified using International Classification of Diseases, Ninth Revision (ICD-9) and Tenth Revision (ICD-10) codes corresponding to underlying causes of death.

### Other Variables

Demographics (age, sex, race), smoking status (current or past), education level, family income, and medication intake were self-reported during an in-home interview. Body mass index (BMI) was calculated from height and weight measurements obtained during a physical examination conducted at a mobile examination center and defined as weight in kilograms divided by height in meters squared (kg/m²). Systolic and diastolic blood pressure were measured while seated, and up to three measurements were averaged. Hypertension was defined as systolic blood pressure ≥140 mmHg, diastolic blood pressure ≥90 mmHg, or the use of antihypertensive medications. Diabetes mellitus was defined as fasting blood glucose levels ≥126 mg/dL or the use of glucose-lowering medications. Dyslipidemia was defined based on total cholesterol levels or use of lipid-lowering medications. Thyroid disease was ascertained by self-report of a physician diagnosis. Total cholesterol, serum creatinine, glucose, and other components in the metabolic panel were measured using laboratory procedures as reported by the National Center for Health Statistics.

### Statistical Analysis

We compared participant characteristics stratified by the presence of SCMI using Chi-square tests for categorical variables. For continuous variables, we first assessed distributional assumptions using Q–Q plots and formal tests of normality. Normally distributed continuous variables are presented as means ± standard deviations (SD) and compared using independent samples t-tests; non-normally distributed continuous variables are presented as medians with interquartile ranges (IQR) and compared using Wilcoxon rank-sum tests. Categorical variables are presented as counts and percentages.

Multivariable logistic regression was used to assess the cross-sectional association between resting heart rate and the presence of SCMI. We modeled heart rate both categorically (bradycardia, normal, tachycardia) and as a continuous variable (per 10-bpm increase). Models were adjusted as follows: Model 1 was adjusted for sociodemographic variables including age, sex, race/ethnicity, education years, and family income. Model 2 was further adjusted for CV risk factors, including smoking status, alcohol intake, use of antihypertensive medications, BMI, systolic blood pressure, diastolic blood pressure, serum creatinine, hypertension, dyslipidemia, diabetes mellitus, and thyroid disease.

For both all-cause and CV mortality we used multivariable Cox proportional hazards models to assess the associations with resting heart rate. Model 1 adjusted for sociodemographic variables. Model 2 was further adjusted for CV risk factors as described above. Model 3 was additionally adjusted for the presence of SCMI to evaluate whether SCMI attenuates the association between heart rate and mortality. The proportional hazards assumption was evaluated using Schoenfeld residuals and visual inspection of log-log survival plots; no significant violations were identified.

To assess the potential explanatory role of SCMI in the relationship between resting heart rate and mortality, we calculated the percentage change in hazard ratios (HR) after adding SCMI to the fully adjusted models using the formula: (HR_Model2 − HR_Model3) / (HR_Model2 − 1) × 100. This approach quantifies the proportion of the excess risk associated with elevated heart rate that is explained by SCMI ^16,17^.

Subgroup analyses were performed to evaluate the consistency of the association between resting heart rate (per 10-bpm increase) and SCMI across clinically relevant subgroups, including age (≤65 vs. >65 years), sex, race/ethnicity, hypertension status, diabetes status, and BMI category (normal, overweight, obese). Interaction terms were included in the fully adjusted models to formally test for effect modification.

All analyses were conducted without incorporating NHANES survey weights; therefore, the reported associations are valid for the analytic sample but should not be interpreted as nationally representative estimates of the U.S. population. Prior methodological work indicates that unweighted regression analyses yield unbiased and often more efficient estimates of exposure–outcome associations when key sampling-related covariates are included in the model ^18–20^. This analytic strategy aligns with approaches used in several previous NHANES III investigations ^21,22^. All statistical analyses were performed using The Jamovi project (2025) (Version 2.4.14.0) Retrieved from https://www.jamovi.org. A two-sided P value of 0.05 was used for hypothesis testing.

## Results

### Baseline Characteristics

The study included 7,152 participants with a mean age of 59.0 ± 13.3 years; 53.4% were women, 72% non-Hispanic whites. The mean resting heart rate was 68.5 bpm; 95.7% of participants had normal heart rate, 3.3% had bradycardia, and 1.0% had tachycardia. SCMI was present in 1,744 participants (24.3%). As compared with participants without SCMI, those with SCMI were older, more likely to be male, and more likely to have diabetes, hypertension, or current smoking. The prevalence of SCMI varied according to heart rate category: 45.1% among participants with tachycardia, 24.4% among those with normal heart rate, and 18.9% among those with bradycardia **(Table 1).**

**Table 1.**
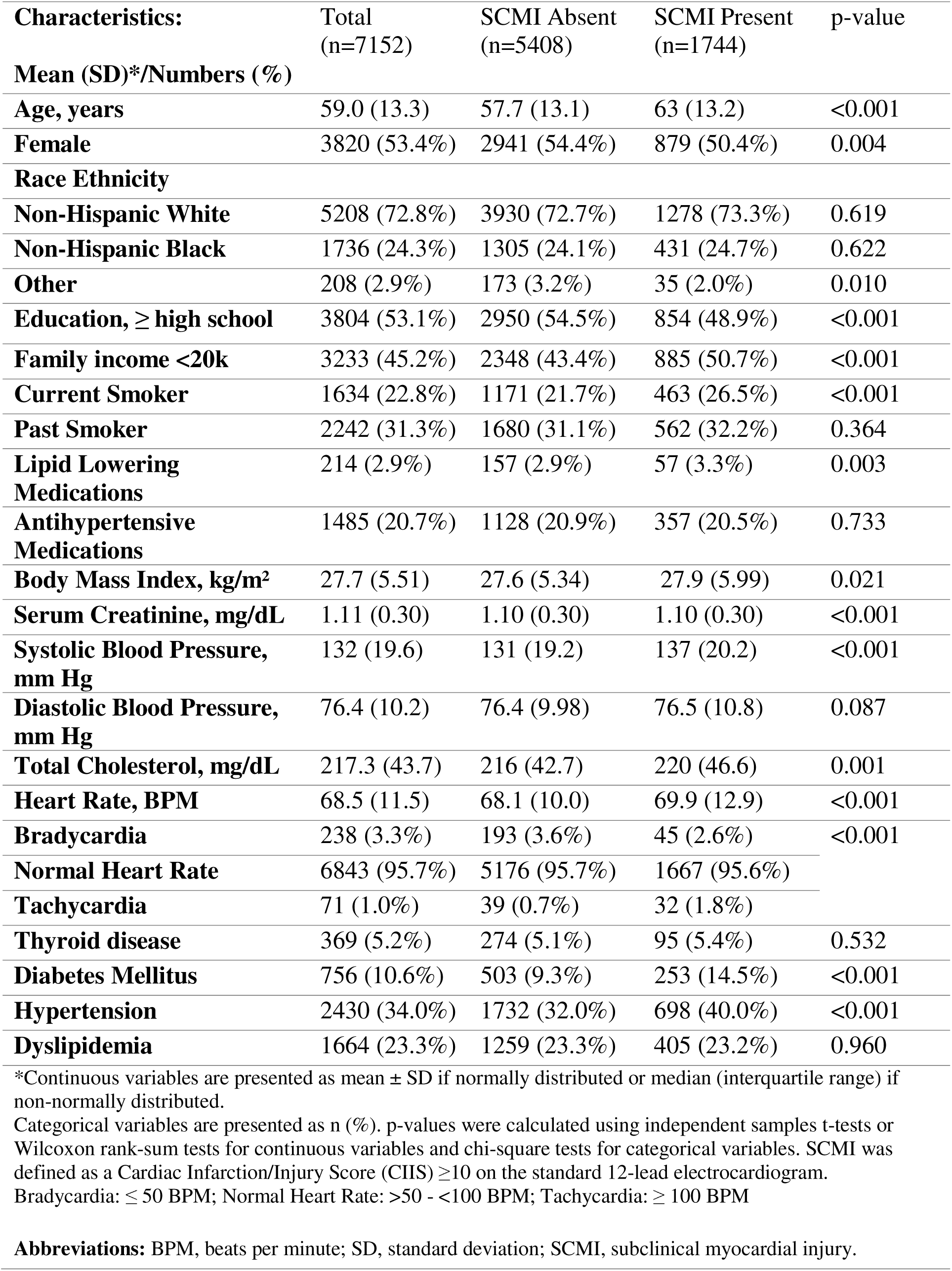
Baseline Characteristics.

### Association Between Resting Heart Rate and SCMI

Tachycardia was associated with more than twice the odds of SCMI as compared with normal heart rate, an association that persisted after adjustment for CV risk factors. Conversely, bradycardia appeared protective against SCMI. The graded nature of this relationship was confirmed when heart rate was modeled continuously: each increase of 10 bpm was associated with a 14% increase in the odds of SCMI, independent of traditional risk factors **(Table 2).**

**Table 2:** Association between Heart Rate Groups and Subclinical Myocardial Infarction.

| HR Group | SCMI N (%) |  | Model 1 |  | Model 2 |  |
| --- | --- | --- | --- | --- | --- | --- |
|  | Present | Absent | OR (95% CI) | p-value | OR (95% CI) | p-value |
| Normal HR | 1667 | 5176 | Ref | -- | Ref | -- |
| Bradycardia | 45 | 193 | 0.6497 (0.46- 0.91) | 0.012 | 0.66 (0.47 - 0.94) | 0.022 |
| Tachycardia | 32 | 39 | 2.45(1.52- 3.96) | <□.001 | 2.34 (1.42-3.88) | <□.001 |
| HR per 10-BPM increase | -- |  | 1.16 (1.11- 1.22) | <□.001 | 1.14(1.08 – 1.19) | <□.001 |

SCMI was defined as a Cardiac Infarction/Injury Score (CIIS) ≥10 on the standard 12-lead electrocardiogram. Model 1 adjusted for age, sex, race/ethnicity, education years, and family income. Model 2 adjusted for Model 1 covariates plus smoking status, alcohol intake, use of antihypertensive medications, body mass index, systolic blood pressure, serum creatinine, dyslipidemia, diabetes mellitus, and thyroid disease. Bradycardia: ≤50 BPM; Normal heart rate: >50–<100 BPM; Tachycardia: ≥100 BPM.
**Abbreviations:** BPM, beats per minute; OR, odds ratio; CI, confidence interval.

### Resting Heart Rate and Mortality Risk

During a median follow-up of 13.9 years, 2,311 participants (32.3%) died, including 933 (13.1%) from CV causes. The mortality risk associated with tachycardia was substantial: participants with tachycardia had more than three times the risk of all-cause death and twice the risk of CV death compared with those with normal heart rate. This relationship was continuous, with each 10-bpm increment conferring incrementally higher risk. In contrast, bradycardia showed no adverse association with mortality and appeared mildly protective **(Table 3**, **Figure 2).**

**Figure 2.**
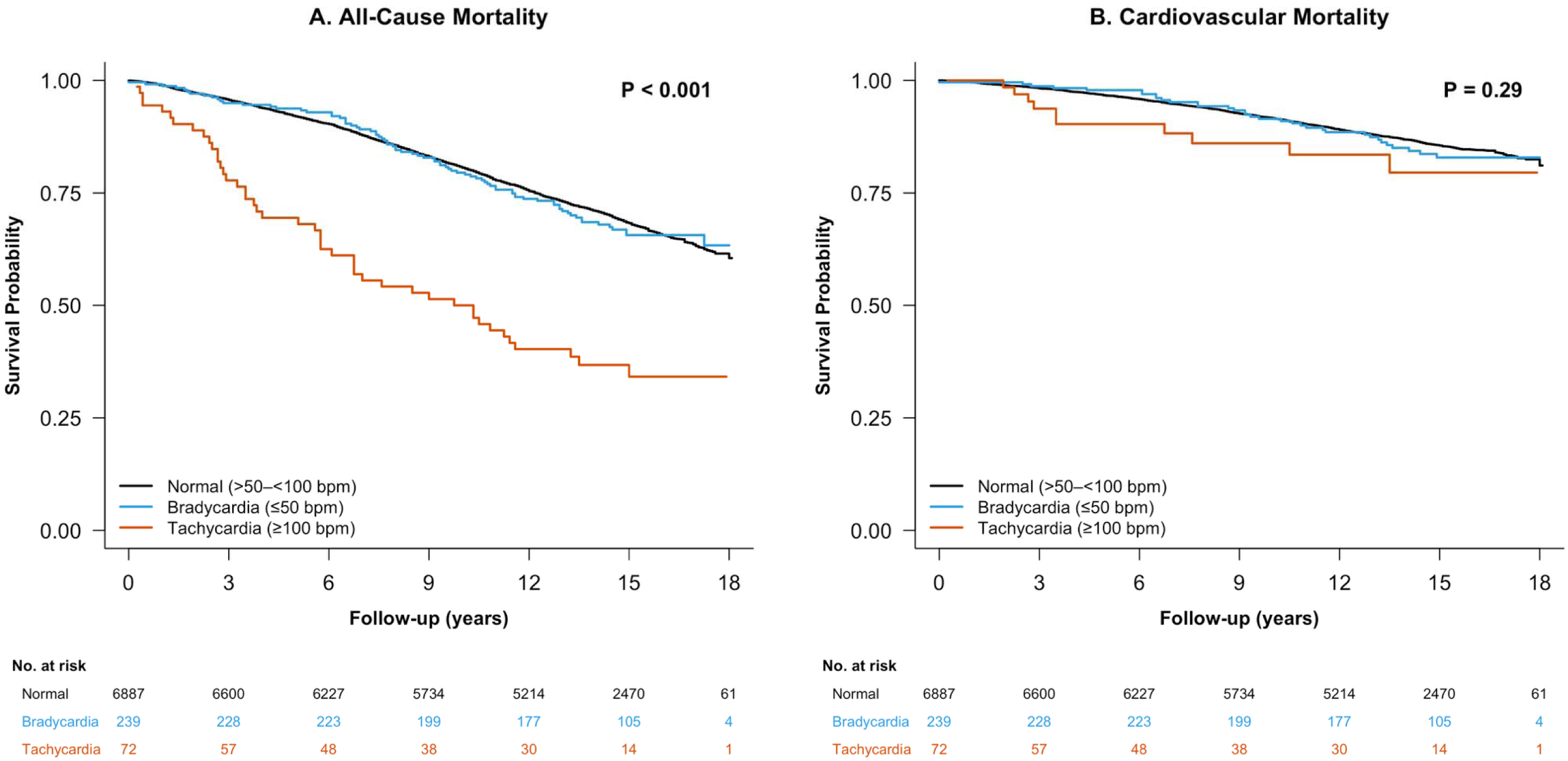
Kaplan-Meier survival curves stratified by resting heart rate categories. Panel A presents all-cause mortality and Panel B presents cardiovascular mortality across three resting heart rate groups: normal (>50–<100 bpm), bradycardia (≤50 bpm), and tachycardia (≥100 bpm). Abbreviation: bpm, beats per minute.

**Table 3:** Association Between Heart rate and Cardiovascular Mortality.

| Outcome | Participants/events | Model 1 |  | Model 2 |  | Model 3 |  |
| --- | --- | --- | --- | --- | --- | --- | --- |
|  |  | HR (95% CI) | p-value | HR (95% CI) | p-value | HR (95% CI) | p-value |
| Normal HR | 6843/889 | Reference | -- | Reference | -- | Reference | -- |
| Bradycardia | 238/34 | 0.85 (0.60-1.19) | 0.342 | 0.81 (0.57-1.17) | 0.265 | 0.86 (0.60–1.23) | 0.416 |
| Tachycardia | 71/10 | 2.38 (1.28-4.46) | 0.006 | 2.05 (1.06-3.79) | 0.033 | 1.96 (1.01-3.80) | 0.046 |
| HR per 10-BPM increase | -- | 1.16 (1.09-1.22) | <0.001 | 1.16 (1.10-1.23) | <0.001 | 1.14 (1.08-1.21) | <0.001 |

Model 1 adjusted for age, sex, race/ethnicity, education years, and family income. Model 2 adjusted for Model 1 covariates plus smoking status, alcohol intake, use of antihypertensive medications, body mass index, systolic blood pressure, serum creatinine, dyslipidemia, diabetes mellitus, and thyroid disease. Model 3 adjusted for Model 2 covariates plus presence of subclinical myocardial injury (SCMI); addition of SCMI accounted for 8.6% attenuation of the heart rate–cardiovascular mortality association. SCMI was defined as a Cardiac Infarction/Injury Score (CIIS) $\geq 10$ on the standard 12-lead electrocardiogram. Bradycardia: $\leq 50$ BPM; Normal heart rate: $>50$ – $<100$ BPM; Tachycardia: $\geq 100$ BPM.
**Abbreviations:** BPM, beats per minute; HR, hazard ratio; CI, confidence interval.

### Attenuation by SCMI

Adjustment for SCMI attenuated the association between elevated heart rate and mortality, suggesting that silent myocardial damage partially explains the prognostic impact of elevated heart rate. The attenuation was similar for all-cause mortality than for CV mortality. SCMI accounted for approximately 5% of the excess all-cause mortality and 8.6% for CV mortality risk **(Table 4**, **Figure 3).**

**Figure 3.**
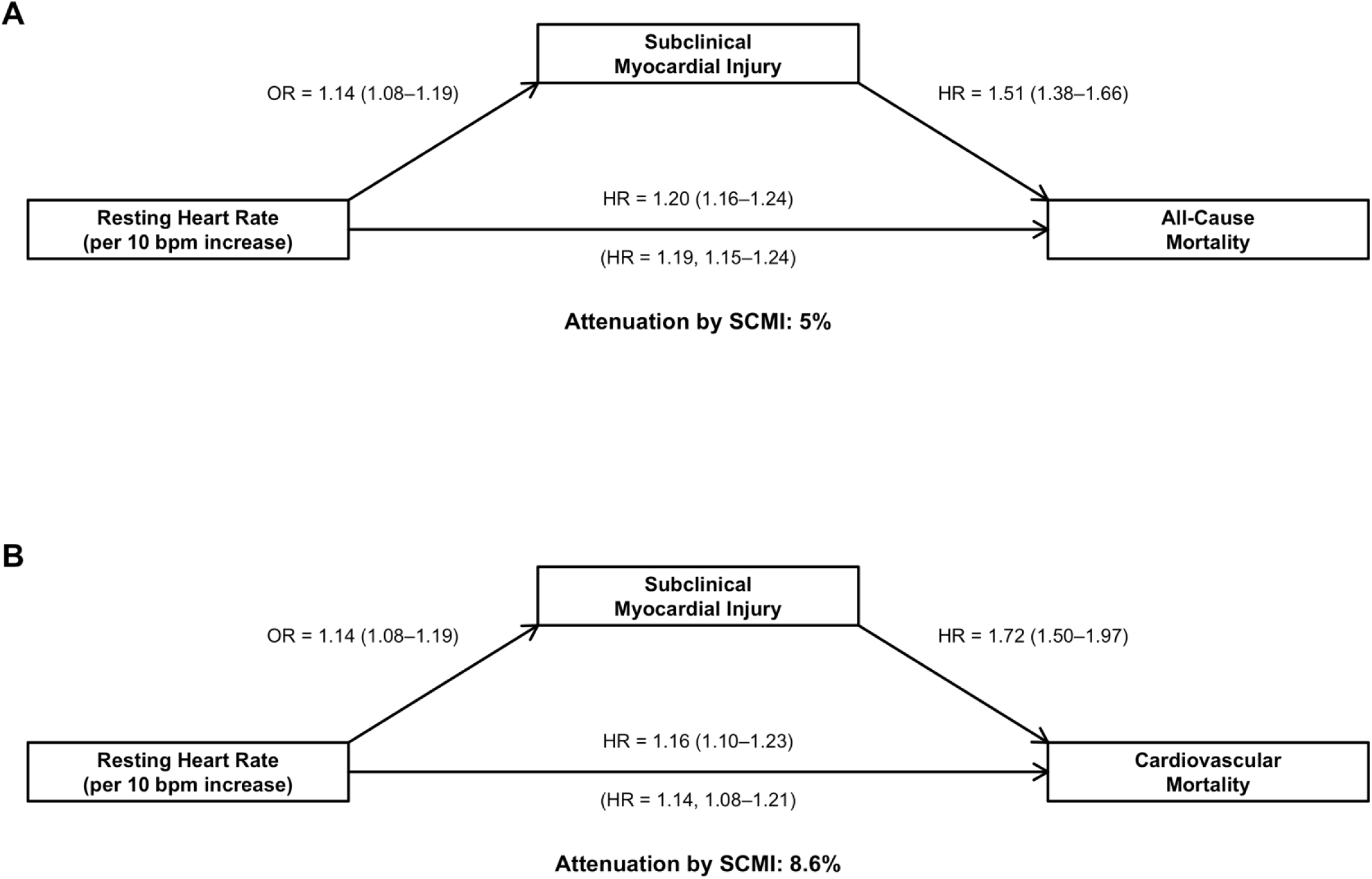
Attenuation of the association between resting heart rate and mortality after adjustment for subclinical myocardial injury. Panel A examines all-cause mortality and Panel B examines cardiovascular mortality, showing direct and indirect pathways through subclinical myocardial injury. Abbreviations: OR, odds ratio; HR, hazard ratio; SCMI, subclinical myocardial injury.

**Table 4.** Association Between Heart rate, SCMI and All-cause Mortality.

| Outcome | Participants/<br>events | Model 1 |  | Model 2 |  | Model 3 |  |
| --- | --- | --- | --- | --- | --- | --- | --- |
|  |  | HR (95% CI) | p-value | HR (95% CI) | p-value | HR (95% CI) | p-value |
| Normal HR | 6843/2186 | Reference | -- | Reference | -- | Reference | -- |
| Bradycardia | 238/79 | 0.80 (0.64-1.01) | 0.057 | 0.78 (0.61-0.99) | 0.040 | 0.79 (0.62-1.01) | 0.057 |
| Tachycardia | 71/46 | 3.54 (2.63-4.76) | <0.001 | 3.58 (2.63-4.88) | <0.001 | 3.45 (2.53-4.69) | <0.001 |
| HR per 10-BPM<br>increase | -- | 1.21 (1.17-1.25) | <0.001 | 1.20 (1.16-1.24) | <0.001 | 1.19 (1.15-1.24) | <0.001 |

Model 1 adjusted for age, sex, race/ethnicity, education years, and family income. Model 2 adjusted for Model 1 covariates plus smoking status, alcohol intake, use of antihypertensive medications, body mass index, systolic blood pressure, serum creatinine, dyslipidemia, diabetes mellitus, and thyroid disease. Model 3 adjusted for Model 2 covariates plus presence of subclinical myocardial injury (SCMI); addition of SCMI accounted for 5% attenuation of the heart rate–all-cause mortality association. SCMI was defined as a Cardiac Infarction/Injury Score (CIIS) $\geq 10$ on the standard 12-lead electrocardiogram. Bradycardia: $\leq 50$ BPM; Normal heart rate: $>50$ – $<100$ BPM; Tachycardia: $\geq 100$ BPM.
**Abbreviations:** BPM, beats per minute; HR, hazard ratio; CI, confidence interval.

### Subgroup Analyses

The association between resting heart rate and SCMI was robust across all prespecified subgroups, with no evidence of effect modification by age, sex, race, hypertension, diabetes, or body-mass index. This consistency supports the robustness of these findings across clinically relevant subgroups (**Table 5).**

**Table 5.** Subgroup Analysis of the Association between Resting Heart Rate and Subclinical Myocardial Injury.

| Subgroup |  | Odds Ratio<br>(95% Confidence Interval) *‡ | Interaction *p*-value<sup>‡</sup> |
| --- | --- | --- | --- |
| Age | ≤ 65 | 1.14 (1.07-1.21) | 0.7030 |
|  | >65 | 1.16 (1.08 - 1.25) |  |
| Gender | *Men* | 1.20 (1.12-1.28) | 0.956 |
|  | *Women* | 1.10(1.03-1.18) |  |
| Race | *White* | 1.19 (1.12-1.26) | 0.131 |
|  | *Black* | 1.08 (0.97-1.19) |  |
|  | *Other* | 1.49(0.99-2.21) |  |
| Hypertension | *Present* | 1.20 (1.13-1.26) | 0.093 |
|  | *Absent* | 1.09 (1.03-1.15) |  |
| Diabetes | *Present* | 1.16 (0.94 - 1.20) | 0.1710 |
|  | *Absent* | 1.09 (0.95 - 1.23) |  |
| BMI | *Normal* | 1.16 (1.08 - 1.25) | 0.5610 |
|  | *Overweight* | 1.14 (1.06 - 1.26) |  |
|  | *Obese* | 1.11 (1.02 - 1.22) |  |

BMI categorized as normal (18.5–24.9 kg/m<sup>2</sup>), overweight (25.0–29.9 kg/m<sup>2</sup>), and obese (≥30.0 kg/m<sup>2</sup>). \*Odds ratios estimated per 10 BPM increase in resting heart rate. ‡Fully adjusted model includes age, sex, race/ethnicity, education years, family income, smoking status, alcohol intake, use of antihypertensive medications, systolic blood pressure, serum creatinine, dyslipidemia, diabetes mellitus, and thyroid disease. SCMI was defined as a Cardiac Infarction/Injury Score (CIIS) ≥10 on the standard 12-lead electrocardiogram.
**Abbreviations:** BMI, body mass index; BPM, beats per minute; OR, odds ratio; CI, confidence interval.

## Discussion

Among participants in NHANES III, a large population-based survey of U.S. adults, with no baseline CVD, elevated resting heart rate was associated with increased prevalence of SCMI and with long-term mortality from all causes and CV causes. Importantly, SCMI partially attenuated the relationship between elevated heart rate and mortality, even after adjustments were made for traditional CV risk factors. This suggests that silent myocardial damage may represent a key mechanistic pathway through which chronically elevated heart rate contributes to adverse outcomes.

Consistent with our findings, prior work has repeatedly linked elevated resting heart rate to overt cardiovascular disease and adverse clinical outcomes; however, far fewer studies have examined whether heart rate relates to subclinical ischemic processes that precede clinical events ^1,2^. Methods used to ascertain subclinical ischemic or atherosclerotic disease vary across cohorts, yet most population-based studies report prevalence estimates similar to ours. The prevalence of SCMI in our NHANES III cohort (24.3%) was comparable to previously reported rates in U.S. and European populations ^8,9^. Likewise, several cohorts employing coronary artery calcium (CAC) scoring as an alternative marker of subclinical disease, including the Heinz Nixdorf Recall Study, the Multi-Ethnic Study of Atherosclerosis (MESA), and large non-Western Korean cohorts, have shown that elevated resting heart rate correlates with greater subclinical atherosclerotic burden ^23–25^. Although these studies rely on CAC rather than ECG-based detection of myocardial injury, they collectively reinforce the concept that higher resting heart rate is consistently linked to early subclinical cardiovascular pathology across diverse populations.

In our cohort, tachycardia was associated with more than two-fold higher odds of SCMI, even after adjusting for multiple cardiovascular risk factors. This aligns with physiologic mechanisms through which elevated resting heart rate may promote subclinical myocardial injury. Higher heart rate increases myocardial oxygen consumption while simultaneously shortening diastole, consequently decreasing coronary perfusion ^5,26^. This mismatch between oxygen supply and demand may promote repetitive episodes of subclinical ischemia ^27^. Over time, such repeated ischemic insults may result in cumulative myocardial damage detectable by ECG-based scoring systems. Additionally, elevated resting heart rate is often considered a marker of autonomic imbalance ^28,29^. Decreased vagal activity has been associated with heart failure in multiple prior studies, and chronic sympathetic activation has been implicated in myocardial fibrosis, adverse ventricular remodeling, and electrical instability ^30,31^. Each of these processes may contribute to the development of SCMI.

Our findings highlight that elevated resting heart rate may be a modifiable contributor to early myocardial injury rather than merely a secondary marker of cardiovascular stress. Although heart-rate–lowering therapies such as β-blockers, non-dihydropyridine calcium channel blockers, and ivabradine are widely used, their potential to slow early atherosclerotic and ischemic processes remains under-leveraged ^32,33^. Growing experimental and clinical insights suggest that heart-rate reduction may favorably influence vascular biology, coronary perfusion, and myocardial oxygen balance; yet no population-based studies have evaluated whether lowering resting heart rate can prevent or attenuate subclinical myocardial injury through heart rate modulation ^34,35^. This gap is particularly relevant given the prognostic importance of SCMI and its demonstrated attenuation of the heart rate–mortality relationship in our cohort.

The association between elevated heart rate and cardiovascular mortality was modestly attenuated by SCMI (8.6%), indicating that heart-rate–dependent cardiovascular risk likely operates through additional pathways beyond CIIS-detectable injury. Some of these mechanisms include prothrombotic states or accelerated atherosclerosis, which are both independently associated with higher resting heart rate and adverse cardiovascular outcomes ^25,36^. Given the modest attenuation by SCMI, further work is needed to identify the predominant non-ECG mechanisms linking elevated resting heart rate to cardiovascular mortality.

The smaller attenuation observed for all cause mortality (5%) similarly indicates that SCMI explains only a limited component of the relationship between elevated resting heart rate and global mortality risk. Rather than acting as the dominant link in this relationship, subclinical myocardial damage may function as marker of reduced physiologic reserve and potentiate a pro-inflammatory state with multisystem consequences ^37,38^. This aligns with prior findings that elevated resting heart rate predicts non-cardiovascular death as strongly as cardiovascular death, suggesting that both heart rate and SCMI may serve as indices of global physiologic compromise ^22^. Consistent with this interpretation, large cohort studies have linked subclinical myocardial injury to increased risk of death from diverse causes, not solely cardiovascular events ^39,40^. Future studies pairing ECG infarction scores with cause-of-death may provide additional insight into which mortality categories are most strongly associated with SCMI.

### Limitations

This study used data from NHANES III, conducted between 1988 and 1994. Changes in CV risk factor prevalence and treatment patterns since that time may limit applicability to contemporary populations. The cross-sectional assessment of heart rate and SCMI at baseline precludes determination of temporal sequence; while we hypothesize that elevated heart rate contributes to the development of SCMI, it is also possible that underlying cardiac abnormalities simultaneously elevate heart rate and predispose to subclinical injury. Resting heart rate was measured at a single time point, which may not fully capture an individual’s typical heart rate or heart rate variability over time. Lastly, because only 71 participants (1%) had tachycardia, our power to detect associations in this subgroup was limited, and the confidence intervals for these estimates were correspondingly wide. It is essential that these findings be confirmed in other populations.

## Conclusion

In this analysis of NHANES III participants, elevated resting heart rate was independently associated with increased odds of subclinical myocardial injury and with higher long-term risk of both all-cause and CV mortality. Importantly, SCMI partially attenuated the association between elevated heart rate and mortality, suggesting that silent myocardial damage may represent one mechanistic pathway through which chronically elevated heart rate contributes to adverse outcomes. These results suggest that assessment of SCMI using ECG-based measures such as the CIIS may enhance CV risk stratification in individuals with elevated resting heart rate. Future research is needed to determine whether interventions that lower resting heart rate can reduce the incidence of SCMI and improve long-term survival.

## Acronym List

BMI: Body mass index
bpm: Beats per minute
CIIS: Cardiac Infarction/Injury Score
CV: Cardiovascular
CVD: Cardiovascular disease
ECG: Electrocardiogram
HR: Hazards ratio
NHANES III: Third National Health and Nutrition Examination Survey
OD: Odds ratio
SCMI: Subclinical myocardial injury

## Author Statement

No conflict of interest existed during the submission process. All authors have approved the manuscript and agreed with submission to the American Journal of Cardiology. We declare that the work described is original research, not previously published, and not considered for publication elsewhere, in whole or in part.

## CRediT authorship contribution statement

Patrick Cheon: Writing – original draft, Writing – review and editing, Visualization.

Mohamed A. Mostafa: Conceptualization, Formal analysis, Data curation, Writing – review and editing.

Mai Z Soliman: Writing – review and editing. Richard Kazibwe: Writing – review and editing.

Elsayed Z Soliman: Conceptualization, Methodology, Supervision, Writing – review and editing, Project administration.

## Data Availability

The data analyzed in this study are publicly available from the National Health and Nutrition Examination Survey (NHANES), conducted by the Centers for Disease Control and Prevention (CDC). NHANES datasets, including laboratory, examination, and questionnaire, can be accessed at https://www.cdc.gov/nchs/nhanes/. Linked mortality follow-up data are available through the National Center for Health Statistics (NCHS) Linked Mortality Files. No new data were created for this study. Derived variables and analytic code used to generate the results are available from the corresponding author upon reasonable request.

## Acknowledgments

The authors thank all team members and participants in the NHANES study.

## Availability of Data and Materials

The datasets generated and/or analyzed during the current study are available on the NHANES’s official website, http://www.cdc.gov/nchs/nhanes.html.

